# SalivaSTAT: Direct-PCR and pooling of saliva samples collected in healthcare and community setting for SARS-CoV-2 mass surveillance

**DOI:** 10.1101/2020.11.23.20236901

**Authors:** Nikhil S Sahajpal, Ashis K Mondal, Sudha Ananth, Allan Njau, Pankaj Ahluwali, Gary Newnam, Adriana Lozoya-Colinas, Nicholas V. Hud, Vamsi Kota, Ted M Ross, Michelle D. Reid, Sadanand Fulzele, Alka Chaubey, Madhuri Hegde, Amyn M Rojiani, Ravindra Kolhe

**Author notes:** Corresponding author Ravindra Kolhe MD, PhD., Medical College of Georgia| Augusta University. BAE 2576, 1120 15th Street| Augusta, GA 30912.

## Abstract

**Background:** The limitations of widespread current COVID-19 diagnostic testing lie at both pre-analytical and analytical stages. Collection of nasopharyngeal swabs is invasive and is associated with exposure risk, high cost, and supply-chain constraints. Additionally, the RNA extraction in the analytical stage is the most significant rate-limiting step in the entire testing process. To alleviate these limitations, we developed a universal saliva processing protocol (SalivaSTAT) that would enable an extraction free RT-PCR test using any of the commercially available RT-PCR kits.

**Methods:** We optimized saliva collection devices, heat-shock treatment and homogenization. The effect of homogenization on saliva samples for extraction-free RT-PCR assay was determined by evaluating samples with and without homogenization and preforming viscosity measurements. Saliva samples (872) previously tested using the FDA-EUA method were reevaluated with the optimized SalivaSTAT protocol using two widely available commercial RT-PCR kits. Further, a five-sample pooling strategy was evaluated as per FDA guidelines using the SalivaSTAT protocol.

**Results:** The saliva collection (done without any media) performed comparable to the FDA-EUA method. The SalivaSTAT protocol was optimized by incubating saliva samples at 95°C for 30-minutes and homogenization, followed by RT-PCR assay. The clinical sample evaluation of 630 saliva samples using the SalivaSTAT protocol with PerkinElmer (600-samples) and CDC (30-samples) RT-PCR assay achieved positive (PPA) and negative percent agreement (NPA) of 95.8% and 100%, respectively. The LoD was established as ∼20-60 copies/ml by absolute quantification. Further, a five-sample pooling evaluation using 250 saliva samples achieved a PPA and NPA of 92% and 100%, respectively.

**Conclusion:** We have optimized an extraction-free direct RT-PCR assay for saliva samples that demonstrated comparable performance to FDA-EUA assay (Extraction and RT-PCR). The SalivaSTAT protocol is a rapid, sensitive, and cost-effective method that can be adopted globally, and has the potential to meet testing needs and may play a significant role in management of the current pandemic.

## Introduction

The emergence of COVID-19 in the city Wuhan, China in December 2019 has rapidly evolved into a pandemic. Since then, severe acute respiratory syndrome coronavirus 2 (SARS-CoV-2) has infected more than 40,997,453 individuals across the globe, and has resulted in at least 1,127,637 COVID-19 related deaths (https://coronavirus.jhu.edu/map.html, last accessed October 21, 2020). The high transmission rate, along with the high percentage of asymptomatic infected individuals, have been identified as the major reason for spread of the disease. Under these circumstances, diagnostic testing for COVID-19 remains the most rationale approach for containing the virus and is of unprecedented importance, because if infected individuals are detected early in the course of their infection, globally implemented strategies such as quarantine and contact tracing can be more effective [1,2].

The diagnostic testing for COVID-19 has relied heavily on nasopharyngeal (NPS) or oropharyngeal swab (OPS) samples collected in universal/viral transport medium (UTM/VTM), followed by RT-PCR based assays that target selected regions of the SARS-CoV-2 *nucleocapsid* (*N*), *envelop* (*E*), *spike* (*S*) and/or *open reading frame* (*ORF*) genes [3] (https://www.centerforhealthsecurity.org/resources/COVID-19/COVID-19-fact-sheets/200410-RT-PCR.pdf, last accessed October 21, 2020). However, the massive global demand for testing has reached crisis level with clearly identifiable regional disparities. The continuation of the first wave of the pandemic in several parts of the world and the risk of a second upsurge in infections in countries with previous decline in cases, highlight the need for a rapid, sensitive, cost-effective, and mass population testing methodology that can be implemented on a global scale [4,5]. The major limitations of the current COVID-19 diagnostic testing regimen lie at both pre-analytical and analytical stages. The pre-analytical variables that influence the performance of the tests pertain primarily to sample type. Although NPS remains the gold standard sample type recommended for COVID-19 diagnostic testing, the collection of NPS samples poses challenges that include exposure risk to healthcare workers, and supply chain constraints pertaining to swabs, transport media and personal protective equipment, with self-collection being difficult and yielding less sensitive results. Furthermore, several reports have highlighted the relatively poor sensitivity of NPS samples in early infection and longitudinal testing [6-8]. The analytical variables that determine the performance of the test are a combination of factors that include the efficiency of RNA extraction, RNA purification, and the sensitivity of the RT-PCR reaction. The RNA extraction and purification have been identified as the major rate-limiting steps in the entire testing protocol, leading to increased turnaround time. Additionally, prolonged turnaround time for results as well as the need for expensive kits, automated instrumentation andtrained personnel, has created additional economic and technological constraints. The scientific community has attempted to eliminate some of these pre-analytical and analytical constraints by utilizing saliva as a sample type and/or performing extraction-free RT-PCR assays, respectively. Several groups have shown comparable or higher sensitivity of saliva compared to NPS samples [9-14]. Although some conflicting reports have been published [15-17], we have previously optimized the processing of saliva samples and demonstrated higher sensitivity of saliva compared to NPS samples in both the healthcare and community setting [18]. Extraction free RT-PCR assay eliminates the major limiting step in the analytic phase of COVID-19 testing. Several groups have demonstrated the feasibility of extraction free RT-PCR reaction maintaining the high sensitivity of the assay with NPS samples [19-22]. It is noteworthy that performing extraction free RT-PCR assay using saliva samples has been found to be a feasible method but only with the following caveats: a) effective for the asymptomatic population; b) requires early morning saliva (pure saliva); and, c) has a limit of detection (LoD) of 6000-12000 copies/ml [23]. Although the study is encouraging, the pre-requisite conditions render it unsuitable for mass population screening, especially because the precise sample collection requirement and the low test sensitivity would lead to a high percentage of false negative results. To address these limitations, we have developed and validated a highly sensitive (limit of detection ∼20-60 copies/ml) extraction free RT-PCR assay (SalivaSTAT) using saliva samples collected in both the healthcare and community setting. The SalivaSTAT protocol enabled us to not only achieve high sensitivity, but also simplified saliva processing, which allowed us to validate a five-sample pooling strategy using the SalivaSTAT-extraction free RT-PCR test (**Figure 1**).

**Figure 1.**
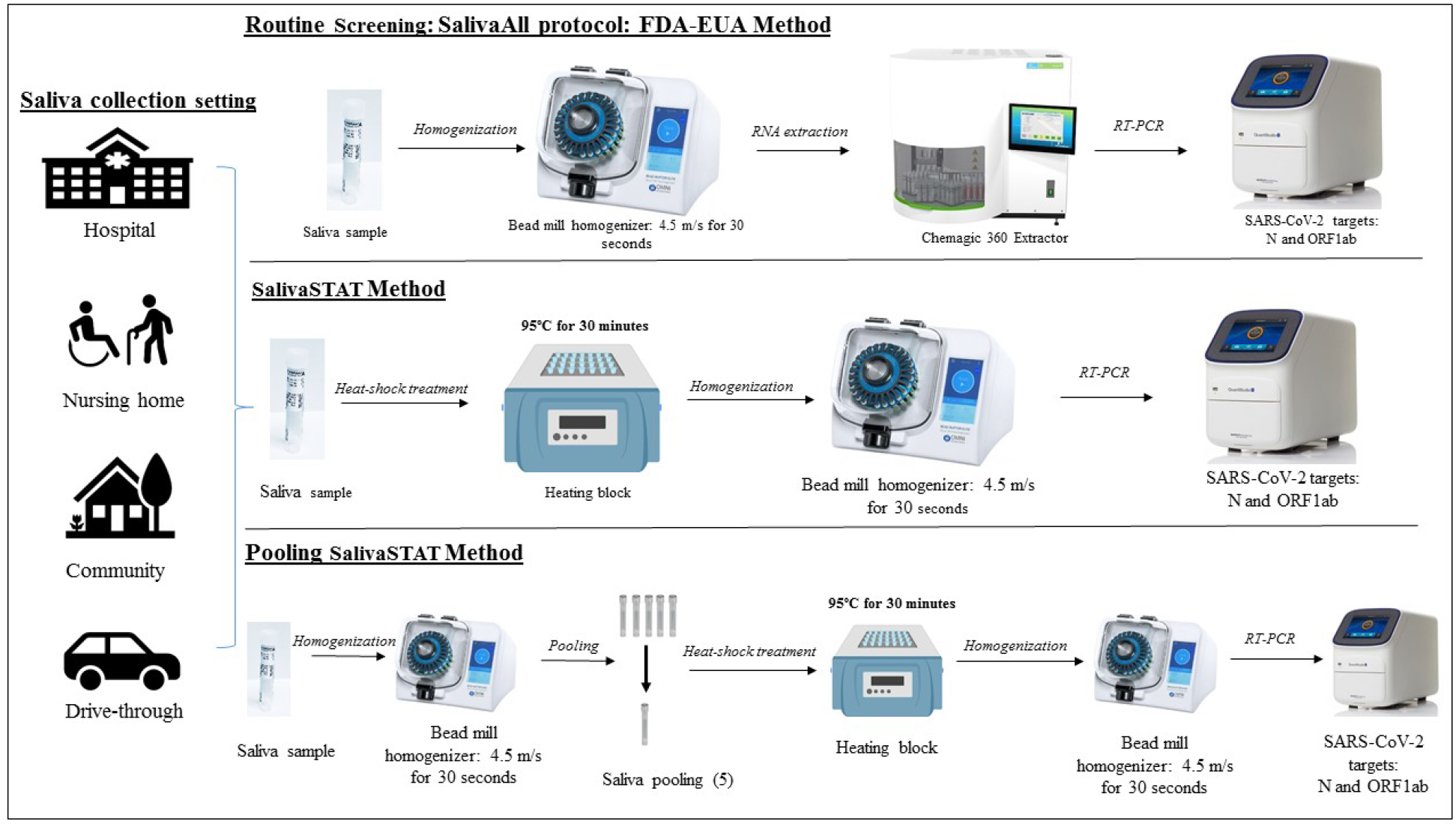
Schematic overview of sample processing and SARAS-CoV-2 assay workflow depicting main steps: Saliva samples collected in healthcare and community setting were tested and validated as follows: Upper panel: Saliva samples processed with SalivaAll protocol for nucleic acid extraction using a semi-automated instrument, followed by RT-PCR for N, ORF1ab gene targets and IC used as extraction and RT-PCR internal control; Middle panel: Saliva samples processed with SalivaSTAT method that included treatment of samples at 95° C for 30 minutes and homogenization using a bead mill homogenizer followed by direct RT-PCR; Lower panel: Saliva samples homogenized using a bead mill before pooling samples with a five-sample pooling strategy followed by SalivaSTAT method for SARS-CoV-2 testing.

## Material and Methods

### Study site and ethics

This single-center diagnostic study was conducted at Augusta University, GA, USA. This site is a CLIA accredited laboratory for high complexity testing and is one of the main SARS-CoV-2 testing centers in the State of Georgia, USA.

### Patient Specimens and setting

The study evaluated 872 saliva samples collected in either healthcare or community setting. Saliva samples were collected in 2 ml vials without any transport media. All samples were stored at 4°C and transported to the SARS-CoV-2 testing facility at Augusta University within 12 hours of collection.

### Assay for the detection of SARS-CoV-2 (FDA-EUA Method)

The assay is based on nucleic acid extraction followed by TaqMan-based RT-PCR assay to conduct in vitro transcription of SARS-CoV-2 RNA, DNA amplification, and fluorescence detection (FDA-EUA assay by PerkinElmer Inc. Waltham, USA). The assay targets specific genomic regions of SARS-CoV-2: *nucleocapsid* (*N*) and *ORF1ab* gene. The TaqMan probes for the two amplicons are labeled with FAM and ROX fluorescent dyes, respectively, to generate target-specific signals. The assay includes an RNA internal control (IC, bacteriophage MS2) to monitor the processes from nucleic acid extraction to fluorescence detection. The IC probe is labeled with VIC fluorescent dye to differentiate its fluorescent signal from SARS-CoV-2 targets.

### Routine Diagnostic Screening (RNA extraction and RT-PCR)

All 872 saliva samples were tested using the FDA-EUA approved assay. In brief, the saliva samples collected in 2ml vials were homogenized at 4.5 m/s for 30 seconds using the Omni bead mill homogenizer (Omni International, USA). An aliquot of 300μl from each sample, positive and negative controls, was then added to respective wells in a 96 well plate. A 5μl internal control (IC), 4μl Poly(A) RNA, 10μl proteinase K and 300μl lysis buffer were then added to each well,. The plate was placed on a semi-automated instrument (Chemagic 360 instrument, PerkinElmer Inc.) following the manufacturer’s protocol. The nucleic acid was extracted in a 96 well plate, with an elution volume of 60μl. From the extraction plate, 10μl of extracted nucleic acid and 5μl of PCR master mix were added to the respective wells in a 96 well PCR plate. The PCR method was set up as per the manufacturer’s protocol on Quantstudio 3 or 5 (ThermoFisher Scientific, USA). The samples were resulted as positive or negative, based on the Ct values specified by the manufacturer.

### Extraction-free RT-PCR assay (SalivaSTAT) optimization

The following parameters were optimized: a) Saliva collection devices, b) Heat-shock treatment and homogenization; c) Heat shock with and without homogenization; d) Saliva sample homogenization.

### Saliva collection devices

Saliva samples were collected in three different collection devices viz. DNA/RNA shield from Zymo Research (cat. no. R1210), Spectrum DNA from Spectrum solutions (cat no. SDNA-1000), and Omni tubes (cat. no. 19-628). The Zymo and Spectrum devices contain transport media that is mixed in 1:1 ratio with saliva, whereas saliva collected in the Omni tubes was media-free. Four previously characterized SARS-Co-V-2 positive samples collected in each device were subjected to 95°C for 10, 20 and 30 minutes respectively, followed by homogenization at 4.5 m/s for 30 seconds using the Omni bead mill homogenizer (Omni International, USA). Following homogenization, the samples were directly processed for RT-PCR using the PerkinElmer RT-PCR kit. The RT-PCR reaction was setup with 10 µl saliva sample and 20 µl reaction master mix (6 µl reagent A, 1.5 µl IC, 1.5 µl reagent B, 1 µl enzyme). The PCR method was set up as per the manufacturer’s protocol on Quantstudio 3 or 5 (ThermoFisher Scientific, USA). The samples were resulted as positive or negative, based on the Ct values specified by the manufacturer.

### Heat-shock treatment and homogenization

Our group and others have previously attempted to optimize the temperature required for direct RT-PCR for NPS samples [19,24]. The next step was to optimize the duration of heat treatment by subjecting four previously characterized positive saliva samples to 95°C for 10, 20 and 30 minutes, respectively, followed by homogenization at 4.5 m/s for 30 seconds using the Omni bead mill homogenizer (Omni International, USA). Subsequently, the samples were directly processed for RT-PCR.

### Heat shock with and without homogenization

Twenty-five saliva samples were subjected to 95°C for 30 minutes, and an aliquot from each sample was either vortexed for 30 seconds or homogenized at 4.5 m/s for 30 seconds using the Omni bead mill homogenizer (Omni International, USA). Following the respective treatment, all samples were directly tested by RT-PCR.

### Saliva sample homogenization

Our group has previously demonstrated the need for homogenization of saliva samples for optimized processing for SARS-Co-V-2 testing. Herein, we perform additional studies to demonstrate that optimal results are achieved using saliva samples for extraction-free RT-PCR using homogenization [18].

### Determination of saliva viscosity

Disposable viscometers were constructed from plastic tubing and plastic transfer pipettes, called Setup A and Setup B, respectively (Figure S1). Standard curves for these viscometers were constructed using water-glycerol solutions and data reported by Segur and Oberstar (25) for the viscosity of water-glycerol mixtures at room temperature ranging from 1 cP (100% water, 0% glycerol) to 1400 cP (0% water, 100% glycerol). Several water-glycerol standards were loaded onto viscometer Setup A for low viscosity liquids (1 cP to 10 cP) and Setup B for higher viscous liquids (100 cP to 1400 cP). For generation of the standard curves, the amount of time required for a specific weight of solvent to flow between two marks on each viscometer was plotted versus the reported viscosity of several water-glycerol mixtures. For Setup A, the viscometer was constructed from plastic tubing with an inner diameter of 1.19 mm and timing marks separated by 240 mm (**Figure S1A**). For Setup B, the viscometer was constructed from a wide bore pipette (Cat # 13-711-6M, Fisher Scientific, Pittsburgh, PA, USA), with the top removed for easy loading, and timing marks separated by 50 mm (**Figure S1B**). Standard curves for both viscometers revealed excellent linear correlations between the measured time for water-glycerol samples to travel between timing marks and the reported viscosities of each mixture (**Figure S2**). The viscosities of saliva samples were measured by loading on either viscometer Setup A or Setup B and measuring the time required for travel between timing marks. The travel times were converted to viscosity measurements by using the standard curves shown in **Figure S2**. We note that our method for viscosity measurement represents an inexpensive and safe (disposable apparatus) adaptation of the Ostwald viscometer (26) which is based on Poiseuille’s law, or Poiseuille’s equation. Briefly, Poiseuille’s equation for viscosity determination can be approximated to: η=Aρt, where η corresponds to the viscosity, A is a constant associated to the viscometer, ρ is the density of the liquid and t is the time the liquid requires to travel a set distance for a given volume of the liquid at a particular temperature. If the Poiseuille equation applies to a solvent, then a plot of η/ρ versus travel time will reveal in linear relationship. The excellent linear correlation of these values for the glycerol-water system confirms the proper functioning of our disposable viscometers (**Figure S1**).

### Weight Distribution

The entire saliva sample was transferred to a pre-weighed 1.5 ml Eppendorf tube, and then weighed again to determine the total weight of the saliva. Samples were centrifuged at 4500x g for 2 min to pellet their high viscosity elements. The low viscous fraction was transferred back to the original tube and the remaining saliva was re-weighed to determine the weight of the high viscosity fraction. The difference between the total weight and the weight of the high viscosity fraction provided the weight used for viscosity measurements.

### SalivaSTAT: Clinical sample evaluation using commercial kits

Six hundred (600) previously tested saliva samples were evaluated using SalivaSTAT protocol and tested with Perkin Elmer Inc. (FDA-EUA) RT-PCR assay, and 30 saliva samples with the CDC RT-PCR assay.

### Pooling saliva samples for Mass Population screening

A five-sample pooling strategy was evaluated as per FDA guidelines (https://www.fda.gov/medical-devices/coronavirus-disease-2019-covid-19-emergency-use-authorizations-medical-devices/vitro-diagnostics-euas). Briefly, 25 previously confirmed positive saliva samples were identified to create 25 positive pools each comprised of one positive and four negative samples. The Ct values of positive samples ranged from (*N*: 19.8-36.8, *ORF1ab*: 25.3 –Undetermined). Similarly, 25 negative sample pools were created comprised of five negative samples. All saliva samples were processed with SalivaSTAT protocol and tested using the PerkinElmer RT-PCR assay.

## Results

### Saliva collection devices

For the saliva samples collected in Zymo, Spectrum and Omni devices, the amplification for IC and SARS-CoV-2 *N* and *ORF1ab* target genes was observed only in saliva samples collected in Omni vials (which were devoid of any media), whereas no amplification was observed in saliva samples collected in Spectrum or Zymo devices. Thus, the process variables for extraction free PCR were optimized using saliva samples collected in Omni devices.

### Heat-shock treatment and homogenization

Four previously tested positive samples were subjected to 95°C for 10, 20 and 30 minutes followed by homogenization and direct RT-PCR. Of the four samples, the Ct values for IC, *N* and *ORF1ab* gene were comparable at all three conditions. However, in samples 3 and 4, the Ct value for *N* and *ORF1ab* gene remained undetermined at 10 minutes treatment, whereas it was comparable at 20 and 30 minutes treatment. (**Figure 2**).

**Figure 2.**
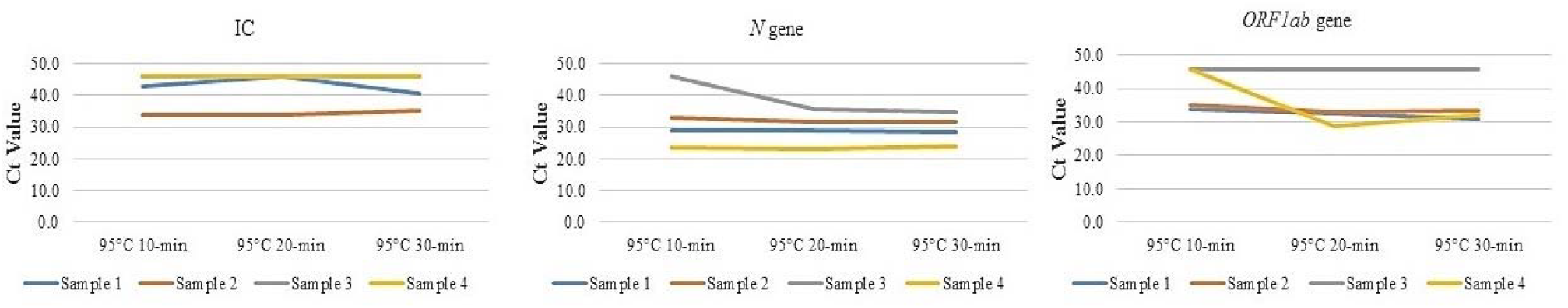
IC, *N* and *ORF1ab* gene Ct values of four previously tested positive samples treated at 95°C for 10, 20 and 30 minutes followed by homogenization and direct RT-PCR.

### Heat-shock treatment with and without homogenization

Twenty-three saliva samples (22 negative and 1 positive) were subjected to 95°C for 30 minutes, and an aliquot from each sample was either vortexed for 30 seconds or homogenized followed by direct RT-PCR. The Ct values for IC (37.12 ± 2.93 vs. 35.03 ± 2.36) were significantly higher in samples that were vortexed compared to homogenization. The amplification curve for the positive sample did not result in an S-shaped curve with vortexing, while a standard amplification curve with comparable Ct value to the FDA-EUA method was identified with the homogenization method. Further, six samples remained invalid with the vortex protocol compared to no invalid samples with the homogenization method (**Figure 3**).

**Figure 3.**
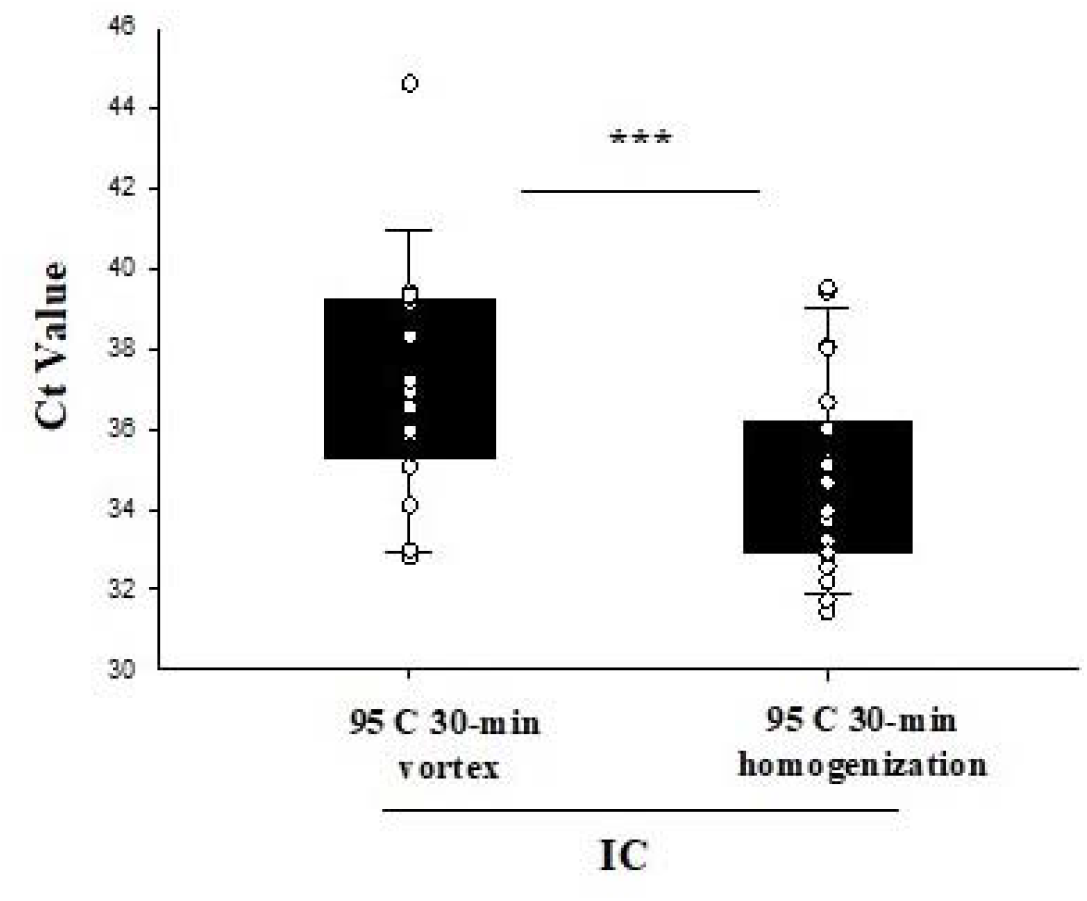
IC Ct values of twenty-two saliva samples subjected to 95°C for 30 minutes, and either vortexed for 30 seconds or homogenized followed by direct RT-PCR.

### Saliva sample homogenization

The effect of homogenization of the saliva samples was evaluated by performing viscosity measurement studies.

### Viscosity Determination

The various saliva samples were loaded onto either of two types of readily-fashioned, disposable viscometers (see Methods for more information) to obtain the time required for each sample to pass between two timing markings (see Supplemental Table 1A and 1B). The average time for each sample was divided by the sample density and these values were compared to the standardized curves **Figure S2A** and **S2B**, to determine the sample viscosity, as shown on Supplemental Table 2. The unprocessed samples had the highest viscosity ranging from 176 cP to 677 cP (between the viscosity of olive oil and honey), compared to processed samples with 2.1 cP to 3.1 cP, which have a viscosity close to the viscosity of water (1 cP).

### Weight Distribution

Saliva samples do not have uniform consistency and vary from watery, thick, sticky, to frothy depending on the amount of proteins in the saliva [27]. For viscosity measurements it was necessary to use a benchtop centrifuge to separate non-flowable material from the flowable material that could be run through a viscometer. To determine the percentage of flowable material that was used for viscosity studie it was necessary to separate and weigh these two phases of the saliva material (see Methods for more information). The inconsistency of the unprocessed samples spanned a range of 61.2% to 98.4% unflowable material that could not be used in viscosity measurements (**S Table 3**).

### SalivaSTAT: Clinical sample evaluation using commercial kits

The SalivaSTAT method was optimized with the following conditions: saliva collected in the media-free Omni tubes was subjected to 95°C for 30 minutes followed by homogenization at 4.5 m/s for 30 seconds using the Omni bead mill homogenizer (Omni International, USA). Following homogenization, the samples were directly tested with RT-PCR assay. Six-hundred (600) saliva samples, comprised of 61 positive and 539 negative samples, were tested with the SalivaSTAT method using the PerkinElmer RT-PCR assay. The Ct values for *N* gene were comparable (29.3 ± 4.8 vs. 28.3 ± 5.6), whereas the Ct value for IC (34.5 ± 3.7 vs. 32.2 ± 1.9) and *ORF1ab* (33.0 ± 4.3 vs. 25.9 ± 5.5) genes were significantly higher with SalivaSTAT compared to FDA-EUA method, respectively. (**Figure 4**).

**Figure 4.**
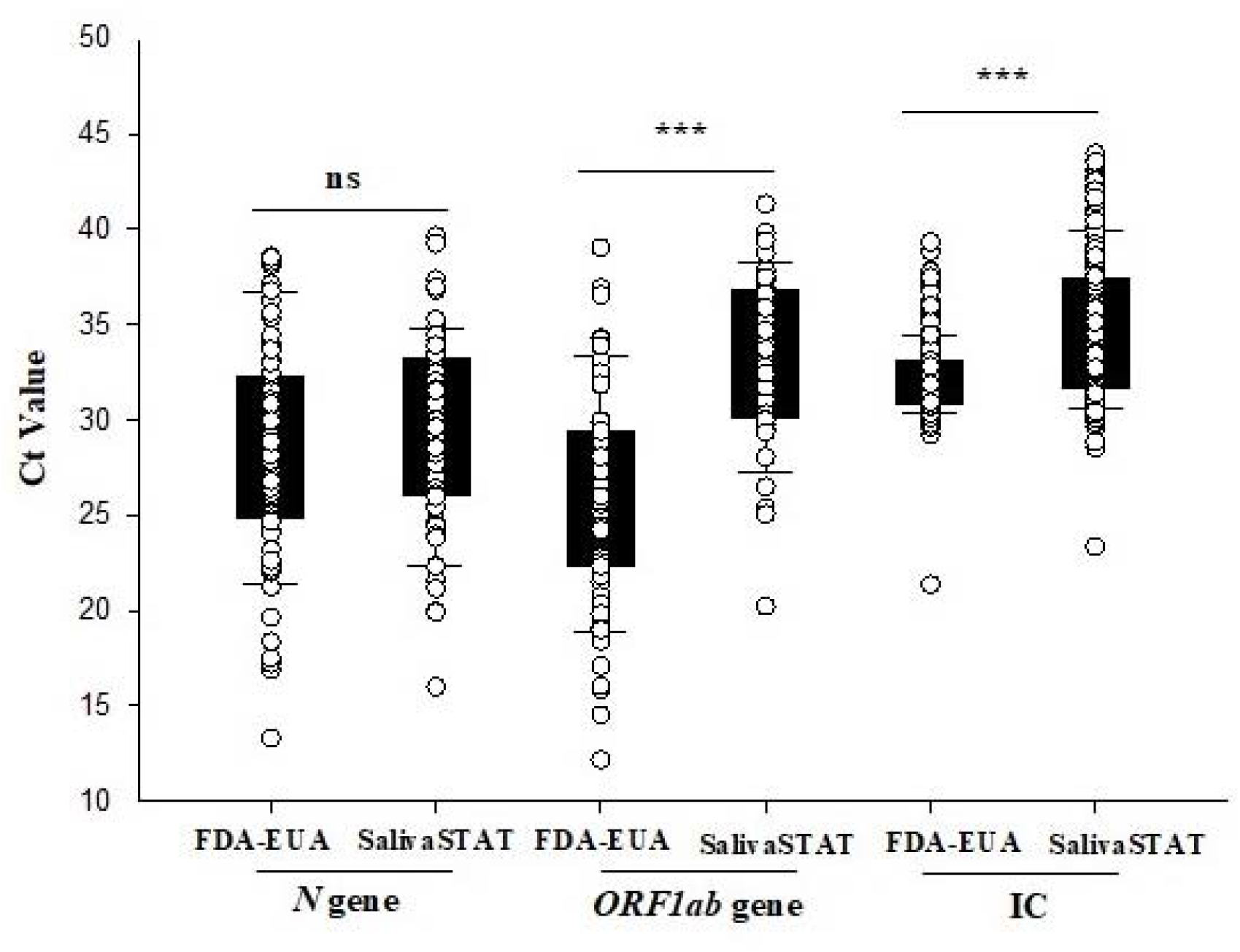
Comparison of Ct values for *N, ORF1ab* genes, and IC of 600 saliva samples evaluated with FDA-EUA (RNA extraction and RT-PCR) method and SalivaSTAT (Direct RT-PCR) method.

Of the 61 positive samples, 95.8% (69/72) were accurately detected by the SalivaSTAT method compared to the FDA-EUA method. The positive samples were selected to represent both strong and weak positives, with Ct values ranging from 16.8-38.5 for the *N* gene, and 14.5-39.0 for *ORF1ab* gene with the FDA-EUA method. Three very weak positive samples (with Ct values of *N*: 38.5, 38.4, 38.2; *ORF1ab*:Und., 36.9, Und., respectively) identified with the FDA-EUA method were not detected with the SalivaSTAT method. Of the 528 negative samples, 498 resulted as negative and 30 as invalid. Similarly, 30 saliva samples, comprised of 16 positive and 14 negative were tested with the SalivaSTAT method using the CDC RT-PCR assay. The Ct values of the 16 positive samples for *N* gene [27.6 ± 5.1 vs. 28.8 ± 4.4 vs. *N1*: 27.5 ± 5.1, *N2*: 29.1 ± 5.0) were found to be comparable with FDA-EUA method and SalivaSTAT-PerkinElmer RT-PCR assay, respectively (**Figure 5**). The overall positive and negative percent agreement was found to be 96% and 100%, respectively. The LOD was determined to be ∼ 20-60 copies/ml by absolute quantification calculation.

**Figure 5.**
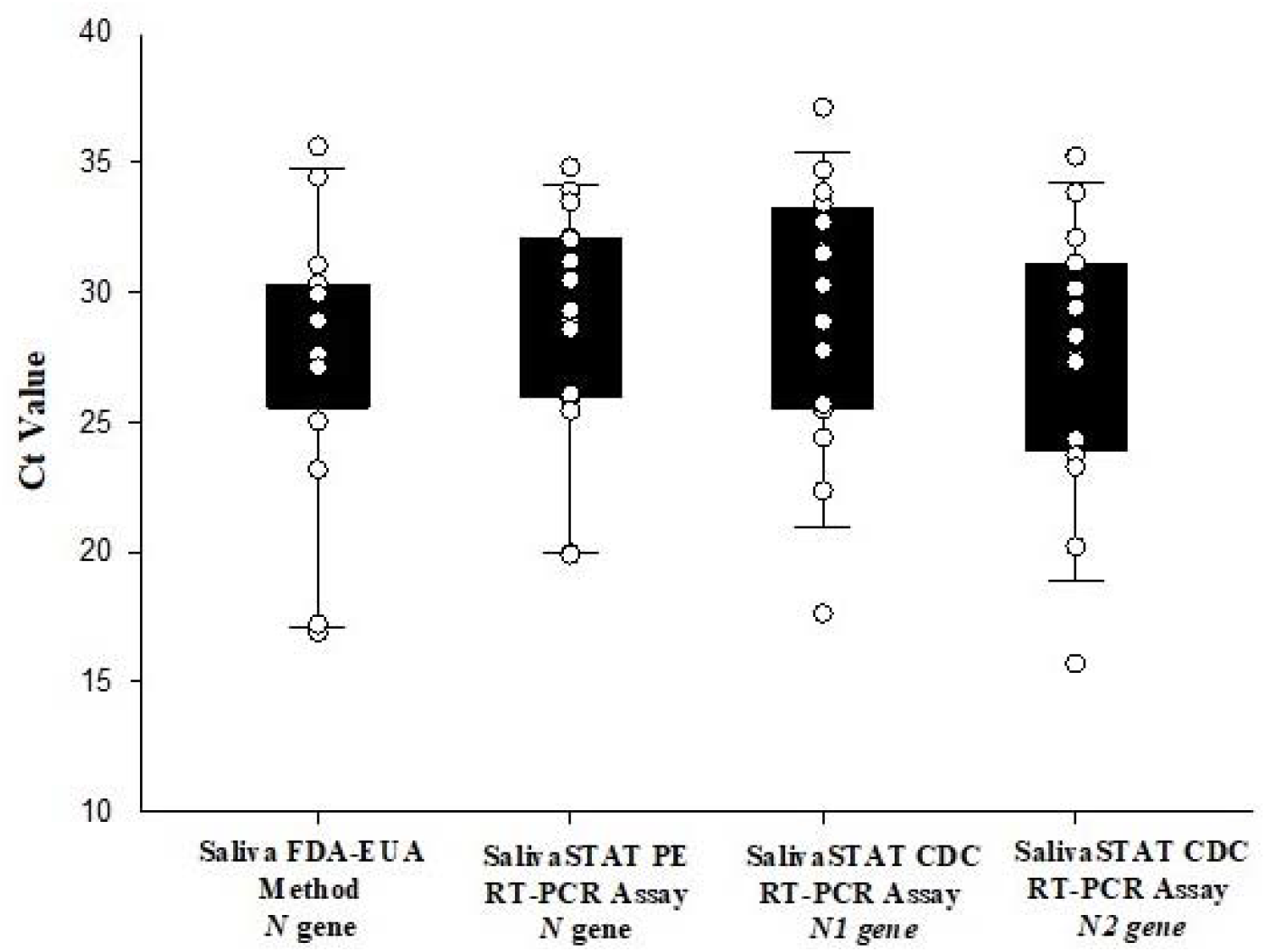
Comparison of *N* gene Ct values of saliva samples evaluated with FDA-EUA method, and SalivaSTAT method using PerkinElmer Inc. (PE) RT-PCR aassay and CDC RT-PCR assay.

### Pooling saliva samples for Mass Population screening

The five-sample pooling strategy was evaluated by comparing the results of the 25 positive and negative pools to individual sample testing results. The pooled testing results demonstrated a 92% positive and 100% negative percent agreement. The *N* and *ORF1ab* gene Ct values were compared between pooled and individual testing. Regression analysis with slope and intercept along with a 95% confidence interval was determined. The shift in Ct value was found to be significant with pooled testing towards higher Ct values, nonetheless, the pools containing positive samples with viral loads close to the assay’s LoD (i.e., weak positives) were accurately detected (**Figure 6**).

**Figure 6.**
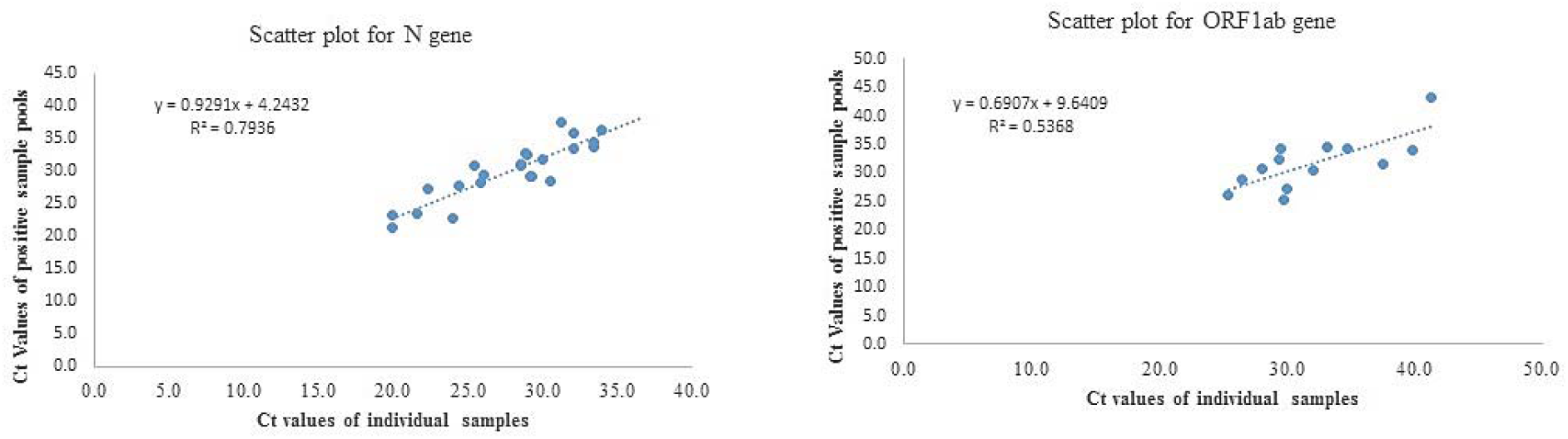
The Ct values comparison of *N* and *ORF1ab* gene with individual testing vs. pool testing.

## Discussion

The COVID-19 pandemic has led to an enormous burden on the health care systems globally, to the point of exhausting currently available resources to manage and/or contain its spread. In this effort, testing for SARS-CoV-2 has been the most critical measure implemented across the globe [1]. The current COVD-19 diagnostic testing regimen primarily relies on NPS samples, followed by qualitative RT-PCR based methods for the detection of SARS-CoV-2. However, several limitations exist in the current methodology at both pre-analytical and analytical stages. In th pre-analytical stage, NPS are associated with exposure risk to healthcare workers, high cost, invasive collection, and supply-chain constraints [6-8]. Further, the RNA extraction step in the analytical stage is the most significant rate-limiting step in this protocol because of wide range of reasons that include, the requirement for competent testing personnel, cost of reagents/ kits, equipment, and turnaround time.

To overcome the pre-analytical and analytical limitations of current COVID-19 testing methods, saliva samples and extraction-free RT-PCR assays have recently been explored. Significant efforts have been made to develop an extraction-free RT-PCR assay using NPS swabs, and several groups have optimized dry swabs, transport media, heat inactivation, different RT-PCR reaction chemistries, and RT-PCR methods [19-22], but minimal information has emerged on extraction-free RT-PCR assay using saliva samples. To our knowledge, only one report has evaluated the performance of extraction-free RT-PCR assay using saliva samples, and this assay was limited to an asymptomatic population, used early morning saliva, and yielded low sensitivity [23]. Hence, the goal of this study was to optimize both pre-analytical and analytical variables by developing a universal saliva processing protocol that would enable an extraction free RT-PCR test using any of the commercially available RT-PCR kits.

In the pre-analytical stage, the most important variables are the collection method and the collection device. Several studies have demonstrated comparable or higher sensitivity of early morning saliva, deep throat saliva, and typical saliva compared to NPS samples, in both the healthcare and community settings [9-14]. Further, most of these studies have used specialized saliva collection devices that mix saliva in 1:1 ratio with a transport media. In the present investigation, saliva samples collected in two specialized collection devices (with media), and one in-house collection device (without media) were evaluated for extraction-free RT-PCR assay. The saliva samples collected in specialized collection devices did not show amplification for either internal control or for the two SARS-CoV-2 *N* and *ORF1ab* gene targets, whereas the saliva samples collected in the in-house collection devices showed amplification for each target with Ct values comparable to the FDA-EUA method. This is consistent with our previous report on extraction-free RT-PCR assay using NPS samples, where NPS samples collected in VTM/UTM did not show amplification for any of the three targets.

The VTM/ UTM appears to inhibit the PCR reaction, and is a consistent observation, as several groups developing the extraction-free RT-PCR assay have either designed their own PCR chemistries, or have validated the input of sample that would allow amplification in their respective RT-PCR methods [19,22]. In addition to being cost-prohibitive and difficult to implement globally, designing alternate PCR chemistries would be challenging in achieving high sensitivity. We therefore attempted to collect saliva samples in the in-house collection device (media-free) which is in alignment with previous report [23].

We and others, have also previously optimized the temperature required for direct RT-PCR for NPS samples [19,24], and thus, our aim was to optimize the duration of the temperature treatment by subjecting four previously characterized positive saliva samples to 95°C for 10, 20 and 30 minutes followed by homogenization. Of the four samples, the Ct values for IC, *N* and *ORF1ab* gene were comparable with all three conditions. However, in sample three and four, the Ct value for *N* and *ORF1ab* gene remained undetermined at 10 min interval, whereas it was comparable at 20 and 30-minutes interval, respectively. Thus, a 30-minute incubation time was deemed optimal for further experiments, as in addition to comparable Ct value results, the 30-minutes interval would inactivate the virus rendering it safe to process in clinical and non-clinical laboratories around the globe. The importance of homogenization of saliva samples after the 30-minute incubation at 95°C, was evident from the significant lower Ct values for IC, *N* and *ORF1ab* targets compared to the samples subjected to vortexing alone.

Homogenization also addresses several critical issues associated with saliva samples. Saliva samples collected in specialized devices or without the use of media has been found to be difficult to pipet by testing personnel, which leads to increased processing time [28]. In addition, the gel-like consistency of saliva samples has led to lower sensitivity and resulted in a higher percentage of invalid results. The saliva samples do not have uniform consistency; varying between watery, thick, sticky, or frothy depending on the amount of constituent proteins. We have previously demonstrated the importance of homogenization of saliva samples, which not only eliminates the processing challenges, but also renders them more sensitive compared to NPS samples [18]. We also evaluated the viscosity of saliva samples before and after homogenization. The unprocessed samples had the highest viscosity ranging from 176 cP to 677 cP compared to the processed samples with 2.1 cP to 3.1 cP, which have a viscosity close to that of water (1 cP). This observation highlights and explains the difficulty these unprocessed samples would pose in accurate pipetting and during the extraction procedure, where uniform mixing of reagents would be challenging. Thus, to eliminate processing challenges and taking cues from our previously published studies that demonstrate the role of homogenization in increasing the sensitivity of saliva samples, we processed each sample with the homogenization step.

The 630-sample clinical evaluation of this optimized extraction-free RT-PCR assay (SalivaSTAT protocol) using two commercial kits, demonstrated an overall positive and negative percent agreement of 96% and 100%, respectively. Interestingly, the Ct value for SARS-CoV-2 *N* gene with SalivaSTAT protocol was comparable to that of the FDA-EUA method. The Ct value for *ORF1ab* and IC were significantly higher with SalivaSTAT compared to FDA-EUA method. These results are in alignment with previously published reports on heat-inactivated direct PCR assay using NPS samples, where comparable Ct values were observed for the N1 gene compared to other targets (E and ORF). Heat treatment cleaves the RNA into short fragments and the best results are obtained with the N1 gene primers, as reported previously [19]. Only three samples that were very weakly positive with a very low viral load of <40 copies/ml were not detected with the SalivaSTAT method. The evaluation using the CDC RT-PCR kit might be deemed more suitable for extraction-free assays, because the assay employs N gene target, and the housekeeping *RnaseP* gene target is extracted in abundance which would lead to zero or minimal invalid results.

In addition to clinical sample evaluation with the SalivaSTAT protocol, we were also able to successfully validate saliva samples with a five-sample pooling strategy. The pooled testing results demonstrated a positive percent agreement of 92% (23/25 pools showing positive results), with two pools that contained the sample with very high Ct value being undetectable. The negative percent agreement was found to be 100%. We have previously demonstrated the feasibility and accuracy of a sample pooling approach with NPS and saliva samples for wide-scale population screening for COVID-19. Herein, we extend the utility and potential benefits of the sample pooling approach for population screening using SalivaSTAT protocol for saliva samples.

Considering the evolving epidemiology of COVID-19 and the reopening of educational and professional institutions, travel, tourism, and social activities, monitoring SARS-CoV-2 will remain a critical public health need for the near future. Therefore, the use of a non-invasive diagnostic test (i.e. saliva collection) and extraction-free RT-PCR methodology will significantly enhance screening and surveillance activities. Taken together, we have optimized an extraction-free direct RT-PCR assay for saliva samples that demonstrated comparable performance to FDA-EUA assay (extraction and RT-PCR). The SalivaSTAT protocol is a rapid, sensitive, and cost-effective method that can be adopted globally, has the potential to accelerate testing needs, and could play a significant role in helping to curb the current pandemic.

## Data Availability

All relevant information/ data is provided in the manuscript or in supplementary file.

## References

1. Taipale J, Romer P, Linnarsson S. Population-scale testing can suppress the spread of COVID-19. medRxiv. 2020.04.27.20078329.

2. Bedford J, Enria D, Giesecke J, Heymann DL, Ihekweazu C, Kobinger G, Lane HC, Memish Z, Oh MD, Schuchat A, Ungchusak K. COVID-19: towards controlling of a pandemic. Lancet. 2020; 395(10229): 1015–8.

3. Patel R, Babady E, Theel ES, Storch GA, Pinsky BA, George KS, Smith TC, Bertuzzi S. Report from the American Society for Microbiology COVID-19 international summit, 23 march 2020: value of diagnostic testing for SARS–CoV-2/COVID-19. mBio. 2020; 10.1128/mBio.00722-20.

4. Xu S, Li Y. Beware of the second wave of COVID-19. Lancet. 2020; 395(10233): 1321–22.

5. Ali I. COVID-19: Are We Ready for the Second Wave? Disaster Med Public Health Prep. 2020; 1–3.

6. Wölfel R, Corman VM, Guggemos W, Seilmaier M, Zange S, Müller MA, Niemeyer D, Jones TC, Vollmar P, Rothe C, Hoelscher M. Virological assessment of hospitalized patients with COVID-2019. Nature. 2020; 581(7809): 465–9.

7. Zou L, Ruan F, Huang M, Liang L, Huang H, Hong Z, Yu J, Kang M, Song Y, Xia J, Guo Q. SARS-CoV-2 viral load in upper respiratory specimens of infected patients. N Engl J Med. 2020; 382(12): 1177–9.

8. Wang W, Xu Y, Gao R, Lu R, Han K, Wu G, Tan W. Detection of SARS-CoV-2 in different types of clinical specimens. JAMA. 2020; 323(18): 1843–4.

9. To KK, Tsang OT, Leung WS, Tam AR, Wu TC, Lung DC, Yip CC, Cai JP, Chan JM, Chik TS, Lau DP. Temporal profiles of viral load in posterior oropharyngeal saliva samples and serum antibody responses during infection by SARS-CoV-2: an observational cohort study. Lancet Infect Dis. 2020; 20(5): 565–574.

10. To KK, Tsang OT, Yip CC, Chan KH, Wu TC, Chan JM, Leung WS, Chik TS, Choi CY, Kandamby DH, Lung DC. Consistent detection of 2019 novel coronavirus in saliva. Clin Infect Dis. 2020; ciaa149. doi:10.1093/cid/ciaa149.

11. Azzi L, Carcano G, Gianfagna F, Grossi P, Dalla Gasperina D, Genoni A, Fasano M, Sessa F, Tettamanti L, Carinci F, Maurino V. Saliva is a reliable tool to detect SARS-CoV-2. J Infect. 2020; 81(1): e45–e50.

12. Yoon JG, Yoon J, Song JY, Yoon SY, Lim CS, Seong H, Noh JY, Cheong HJ, Kim WJ. Clinical Significance of a High SARS-CoV-2 Viral Load in the Saliva. J Korean Med Sci. 2020;35(20):e195.

13. Wyllie AL, Fournier J, Casanovas-Massana A, Campbell M, Tokuyama M, Vijayakumar P, Geng B, Muenker MC, Moore AJ, Vogels CB, Petrone ME. Saliva is more sensitive for SARS-CoV-2 detection in COVID-19 patients than nasopharyngeal swabs. Medrxiv. 2020; https://doi.org/10.1101/2020.04.16.20067835.

14. Rao M, Rashid FA, Sabri FS, Jamil NN, Zain R, Hashim R, Amran F, Kok HT, Samad MA, Ahmad N. Comparing nasopharyngeal swab and early morning saliva for the identification of SARS-CoV-2. Clin Infect Dis. 2020;ciaa1156. doi:10.1093/cid/ciaa1156.

15. Lai CK, Chen Z, Lui G, Ling L, Li T, Wong M, Ng RW, Tso EY, Ho T, Fung KS, Ng ST. Prospective study comparing deep-throat saliva with other respiratory tract specimens in the diagnosis of novel coronavirus disease (COVID-19). J Infect Dis. 2020; jiaa487. doi:10.1093/infdis/jiaa487

16. Jamal AJ, Mohammad M, Coomes E, Powis J, Li A, Paterson A, Anceva-Sami S, Barati S, Crowl G, Faheem A, Farooqi L. Sensitivity of nasopharyngeal swabs and saliva for the detection of severe acute respiratory syndrome coronavirus 2 (SARS-CoV-2). Clin Infect Dis. 2020; ciaa848. doi:10.1093/cid/ciaa848.

17. Becker D, Sandoval E, Amin A, De Hoff P, Leonetti N, Lim YW, Elliott C, Laurent L, Grzymski J, Lu J. Saliva is less sensitive than nasopharyngeal swabs for COVID-19 detection in the community setting. medRxiv, 2020; https://doi.org/10.1101/2020.05.11.20092338

18. Sahajpal NS, Mondal AK, Ananth S, Njau A, Ahluwalia P, Chaubey A, Kota V, Caspary K, Ross TM, Farrell M, Shannon MP. SalivaAll: Clinical validation of a sensitive test for saliva collected in healthcare and community settings with pooling utility for SARS-CoV-2 mass surveillance. medRxiv. 2020; doi: https://doi.org/10.1101/2020.08.26.20182816.

19. Smyrlaki I, Ekman M, Lentini A, Rufino de Sousa N, Papanicolaou N, Vondracek M, Aarum J, Safari H, Muradrasoli S, Rothfuchs AG, Albert J, Högberg B, Reinius B. Massive and rapid COVID-19 testing is feasible by extraction-free SARS-CoV-2 RT-PCR. Nat Commun. 2020; 11(1): 4812. doi: 10.1038/s41467-020-18611-5.

20. U. Kiran, C.G. Gokulan, S.K. Kuncha, D. Vedagiri, K.B. Tallapaka, R.K. Mishra, K. Harshan. Improved and Simplified Diagnosis of Covid-19 using TE Extraction from Dry Swabs. bioRxiv. 2020; https://doi.org/10.1101/2020.05.31.126342.

21. S. Srivatsan, P.D. Han, K.R Van, C.R. Wolf, D.J. McCulloch, A.E Kim, E. Brandstetter, B. Martin, J. Gehring, W. Chen, S. Kosuri. Preliminary support for a “dry swab, extraction free” protocol for SARS-CoV-2 testing via RT-qPCR. bioRxiv. 2020; https://doi.org/10.1101/2020.04.22.056283.

22. S.K. Wee, S.P. Sivalingam, E.P. Yap. Rapid direct nucleic acid amplification test without RNA extraction for SARS-CoV-2 using a portable PCR thermocycler. Genes (Basel). 2020; 11(6): 664.

23. Vogels CB, Watkins AE, Harden CA, Brackney D, Shafer J, Wang J, Caraballo C, Kalinich CC, Ott I, Fauver JR, Kudo E. SalivaDirect: A simplified and flexible platform to enhance SARS-CoV-2 testing capacity. medRxiv. 2020; https://doi.org/10.1101/2020.08.03.20167791.

24. Sahajpal NS, Mondal AK, Njau A, Ananth S, Kothandaraman A, Hegde M, Chaubey A, Padala S, Kota V, Caspary K, Tompkins SM. Clinical validation of innovative, low cost, kit-free, RNA processing protocol for RT-PCR based COVID-19 testing. medRxiv. 2020; https://doi.org/10.1101/2020.07.28.20163626

25. Segur JB, Oberstar HE. Viscosity of glycerol and its aqueous solutions. Ind Eng Chem Res., 1951; 43: 2117–20.

26. Sorrell CA. Liquid viscosity measurement using a buret - instructional technique. J Chem Ed. 1971: 48: 252.

27. Cunha-Cruz J, Scott J, Rothen M, Mancl L, Lawhorn T, Brossel K, Berg J. Salivary characteristics and dental caries: evidence from general dental practices. J Am Dent Assoc. 2013; 144: e31–40.

28. Landry ML, Criscuolo J, Peaper DR. Challenges in use of saliva for detection of SARS-CoV-2 RNA in symptomatic outpatients. J Clin Virol. 2020; 130: 104567. doi:10.1016/j.jcv.2020.104567.

