## Supplemental for "SalivaSTAT: Direct-PCR and pooling of saliva samples collected in healthcare and community setting for SARS-CoV-2 mass surveillance"

Setup A	Setup B

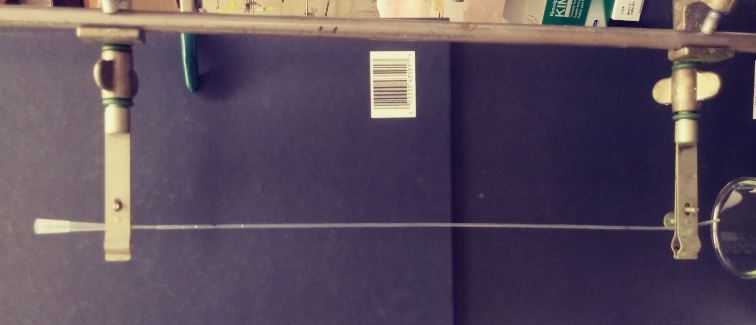

240mm

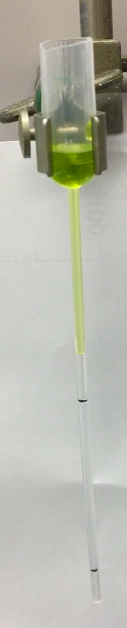

50 mm

**Figure S1: Home-built, disposable viscometers.** Setup A, used for low viscosity samples, consisted of tubing with an inner diameter of 1.19 mm and timing marks separated by 240 mm. Setup B, used for high viscosity samples, consisted of a wide bore pipette (Fisher cat # 13-711-6M), with the top removed for easy loading, and timing marks separated by 50 mm.

Standardized Curves

A	B

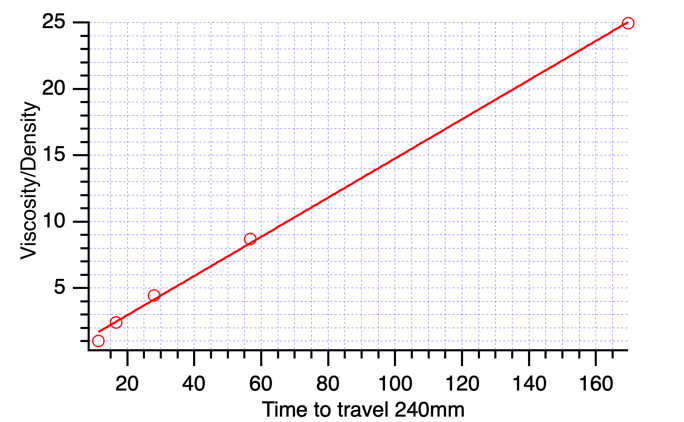

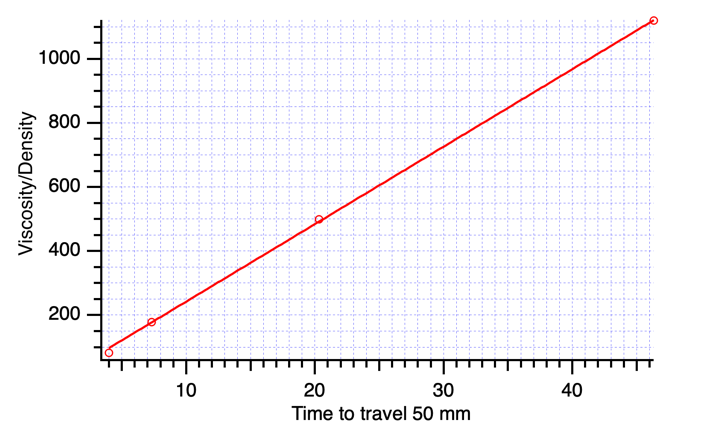

**Figure S2: Standard curves for viscosity determination.** Standardized solutions of glycerol and water were loaded onto a (**A**) long, narrow tube and (**B**) wide bore pipette. Markings on each set were spaced 240mm apart on the long tube and 50mm apart on the pipette. The time was measured for the liquid standards to travel between the markings.

RAW DATA

**Supplemental Table 1: Raw data of saliva samples.** The time required for samples to travel 240 mm (**A**) or 50 mm (**B**) measured in seconds. The processed samples (**A**) were loaded onto viscometer Setup A, while the unprocessed (**B**) were loaded onto viscometer Setup B.

A

| **Homogenized saliva samples** | **112.76** | **242.91** | **194.4** |
| --- | --- | --- | --- |
|  | 19 | 14 | 15 |
|  | 20 | 13 | 15 |
|  | 20 | 13 | 15 |

B

| **Unprocessed Saliva samples** | **112.76** | **242.91** | **194.4** |
| --- | --- | --- | --- |
|  | 8 | 27 | 7 |
|  | 8 | 28 | 7 |
|  | 8 | 26 | 7 |

**Supplemental Table 2: Viscosity of saliva samples.** The viscosity of the saliva samples in centipoise (cP) were determined from the average travel time of the samples between viscometer timing marks (as measured in Suppl Table 1), sample density, and comparison with the standardized curve of the particular viscometer (Suppl Fig 1).

| Setup A |  |  |  |
| --- | --- | --- | --- |
| **Processed Sample** | **Average time for 240mm (s)** | **Density (g/mL)** | **Viscosity (cP)** |
| **112.76** | 19.67 | 1.067 | 3.10 |
| **242.91** | 13.33 | 1.053 | 2.07 |
| **194.4** | 15 | 1.112 | 2.46 |
| Setup B |  |  |  |
| **Unprocessed Sample** | **Average time for 50mm (s)** | **Density (g/mL)** | **Viscosity (cP)** |
| **112.76** | 8 | 1.014 | 196.23 |
| **242.91** | 26 | 1.077 | 677.37 |
| **194.4** | 7 | 1.045 | 176.95 |

Weight Distribution

**Supplemental Table 3: Weight Distribution.** Unprocessed samples were weighed on an analytical balance (mg) to determine the total weight. Material with a low enough viscosity to enter a plastic pipette was weighed (Measured viscous material). The weight of the higher viscosity material was calculated as the difference between the weight of measured viscous material from the total weight. The Percent of useable saliva was calculated by dividing measured viscous material by total weight.

| Unprocessed sample | 112.76 | 242.91 | 194.4 |
| --- | --- | --- | --- |
| Total weight | 422.96 | 719.41 | 1021.57 |
| Measured viscous material | 416.31 | 586.01 | 625.54 |
| High viscous material | 6.65 | 133.4 | 396.03 |
| Percent of useable saliva | 98.4 | 81.5 | 61.2 |

.
